# Applying the COM-B behaviour change model in social and behaviour change message development towards increased uptake of Perennial Malaria Chemoprevention (PMC) delivered through routine immunization platform in Osun State, Nigeria

**DOI:** 10.64898/2026.04.04.26350153

**Authors:** Chinazo Ujuju, Helen Ekpo, Adekemi Anike Ajayi, Helen Hawkings, Daudi Ochieng, Abubakar Ahmed Magaji, Semiu Adebayo Rahman, Ugomma Mong J. Nyananyo, Michael Ekholuenetale, Mary Abosede Adekola, Benjamin Bukky Ilesanmi, Tolu Yetunde Kuye, Titus Kolawole Ojewunmi, Akeem Babatunde Bello, Nnenna Ogbulafor, Rufai-Ahmed Garba, Charles Nzelu, Kolawole Maxwell, Olusola Oresanya, James K. Tibenderana

## Abstract

**Background:** To influence malaria-related behaviours, it is important to understand key behavioural drivers, encourage enablers and address barriers to individuals’ and communities’ adoption of interventions to prevent malaria. The capability(C), opportunity(O), and motivation(M) Behaviour(B) model (COM-B model) was used to inform development of perennial malaria chemoprevention (PMC) social, and behaviour change (SBC) message delivered through routine immunization (RI) platform. This paper presents how the COM-B model was used for designing the SBC messages for PMC using the findings from a qualitative study.

**Methodology:** The COM-B model provided the theoretical framework for designing the PMC SBC intervention by identifying, capability, opportunity motivation for PMC as well as the barriers, and possible enablers for PMC uptake. A qualitative study was conducted as key source of information. Twelve focus group discussions (FGDs) were conducted with the target audience comprising of mothers of children under two years, pregnant women, men, ward development committee members, community mobilizers and health workers. A total of 120 people participated in the study. An SBC workshop was conducted to develop key messages and content for a community dialogue flipbook and facilitators’ guide.

**Results:** Knowledge of malaria signs that prompt mothers to seek health care for their children as well as awareness about malaria prevalence and severity, were identified as capabilities that could drive behaviour change, while forgetting the time to visit the health facility was noted as a hindrance. Opportunities and social influencers included spousal support, the positive influence of health workers, accessibility and affordability of the intervention, and the availability of transportation. Motivation was shaped by the perceived seriousness of malaria as a health problem that could lead to the death of children. Fathers were motivated when they observed reduced malaria burden and improved child health, although a lack of perceived urgency remained a demotivating factor for seeking care. Mothers’ motivation was strengthened by encouragement from husbands, community mobilisers and health workers.

**Conclusion:** The COM-B model provided an effective framework for identifying and developing key messages that informed changes needed to improve capability, opportunities, motivation of individuals and communities towards increased uptake of PMC during PMC pilot study in Osun State Nigeria.

## Background

Malaria remains one of the public health concerns leading to high morbidity and mortality despite being a preventable and treatable disease. Nigeria alone contributes 24.3% of the global malaria cases and 30.3% of the global malaria deaths[1]. Children under five years old and pregnant women being the most vulnerable[2,3]. With high malaria burden in Nigeria, concerted efforts to reduce malaria morbidity and mortality in children is imperative[1]. Perennial Malaria Prevention (PMC) formerly known as Intermittent Preventive Treatment in infants (IPTi) is the administration of preventive antimalarial drugs such as sulfadoxine pyrimethamine (SP) to children to provide personal protection against malaria in areas where transmission rates are moderate to high[4]. The World Health Organization (WHO) recommended the routine immunization (RI) platform for PMC service delivery [4]. Hence PMC complements other preventive malaria control interventions including insecticide treated nets (ITNs) seasonal malaria chemoprevention (SMC), intermittent preventive treatment in pregnancy (IPTp) as well as environmental management in an effort to eliminate malaria in communities with high malaria cases and deaths [5,6].

Uptake of malaria interventions is strongly influenced by human behaviour. Lack of adequate knowledge about malaria transmission and prevention, and the benefits of interventions has resulted to low uptake of malaria interventions. Similarly, perception of malaria and susceptibility to infection influences adoption of prevention behaviour. Culture and social norms such as community beliefs, gender dynamics, and household decision-making affect health seeking behaviour as well as malaria intervention uptake[7–9]. Behaviour change communication which is the use of effective communication to encourage individuals and communities to adopt healthier lifestyles is one of the primary control interventions for malaria which targets humans[10]. The benefits of social behaviour change communication (SBCC) on the uptake of other malaria interventions are well documented [11,12].

Perennial malaria chemoprevention effectiveness (PMC-Effect) study is a cluster randomized control trial designed to provide evidence on the effectiveness and operational feasibility of PMC. Children less than 24 months were offered six doses of SP in the intervention arm at 10 weeks, 14 weeks, six months, nine months, 12 months and 15 months of age, concurrently with EPI vaccinations — the pentavalent vaccine (for diphtheria, tetanus, pertussis, hepatitis B and Haemophilus influenzae type B), vitamin A and measles — where appropriate. Beyond these EPI-linked doses, children receive additional monthly doses between and after the EPI-linked visits up until the child reaches 18 months of age. PMC delivery was from August 2023 to August 2025 in eight Local Government Areas (LGAs) in Osun State, Nigeria[13–15]. To increase the uptake of PMC during the pilot study, social behaviour changing (SBC) and a community-based approach was deployed. Firstly, identifying barriers and understanding enablers of PMC uptake at the individual and community level, and mitigating misconceptions about PMC.

The PMC SBC strategy was developed to guide the design for PMC SBC intervention activities, establish communication objectives and identify the intended audiences to be targeted for PMC SBC activities. To ensure that culturally sensitive messages that are clear, easy to understand, and able to motivate people to take the promoted actions for PMC, the COM-B model [16] and the health belief model [17] informed the SBCC approach. According to Michie et al. (2011) capability (C), opportunity (O), and motivation (M) are three key factors required to change behaviour(B). **Capability** refers to an individual’s psychological and physical ability to participate in an activity[16]. One’s capability is enhanced by knowledge and building skills to practice the desired behaviour. **Opportunity** refers to external factors or social factors outside the individual that could make behaviour possible. While **motivation** refers to the conscious and unconscious cognitive processes that direct and inspire behaviour such as beliefs about capabilities, consequences, reinforcement and emotions.

Perceived susceptibility, benefits, and cue to action are three key components of HBM, a theoretical framework that explains how individuals make decisions about their health. According to the HBM model, individual’s capability to take action to improve their health and well-being depends on their:

1. Perceived susceptibility: The belief that one is at risk of developing a health problem or experiencing a negative health outcome. When individuals perceive themselves as susceptible, they are more likely to take action to prevent or mitigate the risk.
2. Perceived Benefits: The belief that acting will lead to positive health outcomes or benefits. When individuals perceive benefits to taking action, they are more likely to be motivated to do so.
3. Cue to Action: A trigger or prompt that encourages individuals to take action. Cues can be internal (e.g., symptoms) or external (e.g., advice from a healthcare provider, media campaigns).

When these components are present, individuals are more likely to feel motivated to take action, believe they have the ability to take action (self-efficacy), overcome barriers, obstacles and take preventive measures or seek treatment. By improving perceived susceptibility, perceived benefits, and cue to action, individuals are more likely to seek preventive services such as PMC for preventing malaria in children under two years old.

The COM-B model was used to identify behaviours related to malaria and to analyse them in terms of capability, opportunity, and motivation to inform targeted behaviour change messages[18]. This approach enabled a structured assessment of barriers and facilitators and their alignment with appropriate behaviour change techniques. By extending analysis beyond surface-level considerations, the COM-B framework supported the development of messages that were actionable, context-specific, and responsive to behavioural drivers[19]. For designing SBC messages, the model provided a basis for understanding key influences on decision-making and behaviour to enhance uptake of PMC during RI. Widely applied to support behaviour change across diverse health contexts (10,11), COM-B offered a robust structure for determining the changes required to strengthen individual and community capability, opportunity, and motivation to improve PMC.

## Methods

The study was conducted in two randomly selected local government areas (LGAs) - Ede and Obokun selected from the eight LGAs where PMC intervention is being implemented in Osun State Nigeria. The location of the study LGAs is presented in figure 1. Following ethical approval from Osun State Health Research Ethics Committee, (Approval number OSHREC/PRS/569T/591) data collection was conducted from 23^rd^ to 25^th^ July 2024. Participation in the study was voluntary, and participants were informed of their right to withdraw at any stage of the study. The informed consent form was read and explained to all the participants. Questions asked by the participants about the study were addressed before participants signed or thumb printed on the consent forms to confirm their participation in the study.

**Figure 1.**
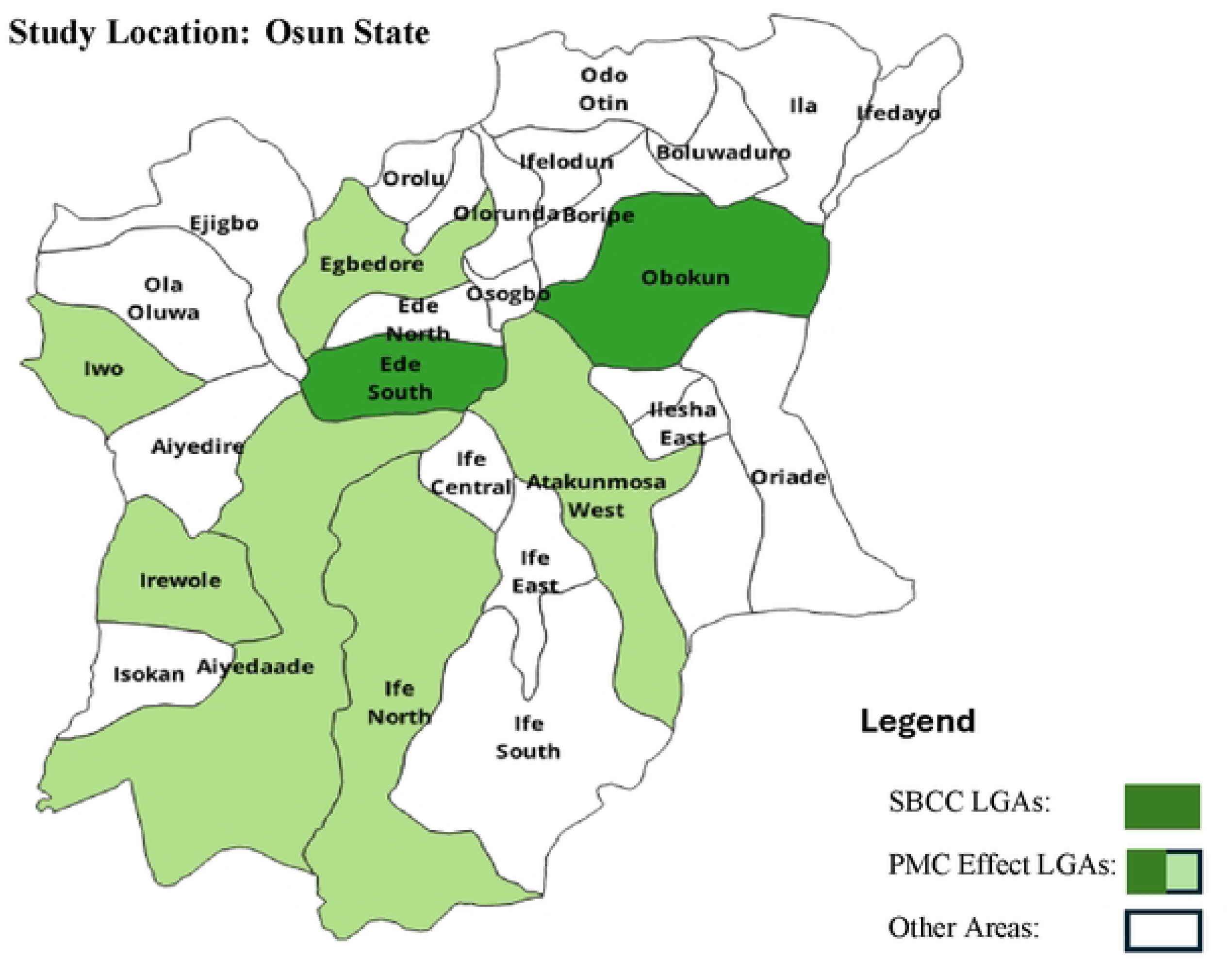
Map of Osun State showing Ede and Obokun LGAs

To obtain a purposive representative sample, 12 focus group discussion (FGD) were conducted with the target audience comprising of 20 mothers of children under two years, 20 pregnant women, 20 men, 20 community mobilizers, 20 ward development committee (WDC) members and 20 health workers. Each focus group had 10 participants and were conducted simultaneously in the two LGAs for three days. A total of 120 persons participated in the FGDs. The FGDs were led by trained moderator and notetakers and the discussions recorded. A discussion guide was developed in English to facilitate discussion with the participants. The guide was pre-tested during the training of moderators and notetakers. The discussions were recorded for ease in data analysis. Most of the discussions conducted in the local dialect while few were in English language. Discussions in the local language were transcribed and translated to English language before transcript-based analysis was conducted. Personal identifiers such as name of participants were not included in the notes and transcripts. Consent was sought and obtained from the participants to record the session for ease of transcription and analysis. Transcription and analysis were done at two levels. First level analysis started in the communities through the noticing, collecting and thinking using the discussion guide, the note taken and the summary template. In the second stage, thematic analysis was conducted using the COMB-Model model as a theoretical framework.

After the barriers and possible enablers for PMC uptake were identified, a three-day SBC workshop was conducted to support the development key messages and content for a community dialogue flipbook and facilitators’ guide focusing on PMC linked to RI. Workshop aimed to enable participants to develop clear and culturally appropriate communication messages, propose suitable graphics for each section of the flipchart, and agree on the next steps for implementing community dialogues in the intervention sites. Participants reviewed and referenced several existing materials, including findings from previous studies on PMC, the PMC SBC strategy, the creative brief, and the recent post implementation study report. These resources were used to analyze motivations and barriers influencing community actions to protect children from malaria, guided by the COM-B behavioural model.

During the first day of the workshop, participants identified and agreed on the key target audiences for the community dialogue sessions. The primary target audience included mothers or caregivers of children under two years of age, pregnant women, Ward Development Committees (WDC), and traditional birth attendants (TBAs). Secondary audiences comprised fathers, healthcare providers, grandparents, heads of households, and community leaders and influencers such as traditional and religious leaders. Tertiary audiences included policymakers and opinion leaders. Using insights from the FGD findings, participants identified several key issues affecting malaria prevention. These included poor environmental sanitation, gaps in knowledge about the severity of malaria, misuse of insecticide-treated nets (ITNs), low utilization of routine immunization services, limited awareness of PMC benefits among older community members, long waiting times at health facilities, restrictions from husbands preventing women from accessing PMC, and inadequate health workforce capacity.

On the second day, participants developed targeted communication messages for each audience group using a structured framework that examined current behaviors, desired behaviors, current actions, desired actions, key messages, and benefit statements. Group presentations were followed by plenary discussions. Participants also identified three to five priority messages per theme, proposed discussion questions for facilitators, and recommended culturally appropriate illustrations for the flipchart, working closely with a graphic artist to conceptualize the visual elements. On the final day, participants refined their outputs based on feedback and existing research materials to ensure the messages were action-oriented and emotionally engaging. They agreed on the community dialogue title, “**PMC is the gateway for child healthy growth and thriving**.” Draft graphics were reviewed and improved, and all group outputs were finalized and submitted for compilation into the community dialogue flipchart and facilitator guide.

### COM-B Model

We followed a structured four-step approach:

1. **Identifying Behavioural Challenges:** Using the COM-B framework, we categorized barriers and enablers identified from the FGDs into capability, opportunity, and motivation domains. For example, limited knowledge of malaria and PMC schedules was categorized as a capability gap, long waiting times and lack of transportation as opportunity gaps, and lack of perceived urgency as motivational gaps.
2. **Defining Target Behaviours:** We used COM-B to specify desirable behaviours among key actors—e.g., mothers consistently bringing children for monthly PMC doses, men supporting attendance, health workers delivering PMC with empathy, and community mobilizers reinforcing correct practices.
3. **Mapping to COM-B Constructs:** We aligned the target behaviours with specific COM-B domains to determine what needed to change. For example:

1. To enhance **capability**, we focused on simplifying messages about knowledge of PMC, knowledge of malaria, prevalence and seriousness.
2. To improve **opportunity**, we promoted social support structures (e.g., husbands providing transport, community leaders endorsing PMC).
3. To boost **motivation**, we used testimonies and reminders (e.g. health workers reminders and caregiver quotes) that emotionally connected caregivers to the protective value of PMC.

### Designing SBC Messages and Delivery Channels

Using the COM-B model enabled us to develop SBC materials (Community dialogue flipbook and discussion guide for facilitators) that resonated with the audience, increasing the likelihood of successful behavior change and improved health outcomes. By linking the findings to the COM-B domains, we tailored messages to address specific barriers.

### Reflexivity

The study was led by a public health expert and the study team consisted of qualitative researchers with experience in malaria programme implementation in Nigeria. Some team members had previously collaborated with the State Ministry of Health, which facilitated access to caregivers, health workers and community leaders and participants willingness to share critical perspectives. To minimize bias, interviews were conducted by trained researchers. The transcripts were also reviewed with the recording by the public health expert prior to analysis and discrepancies were resolved through discussion to ensure balanced interpretation of findings.

## Results

### Capability- Knowledge about malaria, prevalence and seriousness of the disease

Knowledge about the cause of malaria, the symptoms and how the infection could be prevented were mentioned by participants. Signed and symptoms mentioned included high body temperature, nausea, dullness of the eye, loss of appetite, child no longer play actively, anaemia and convulsions. One of the hindrances for capability mentioned by the participants was forgetting the time to visit the health facility. The following quotes buttress their views:

> *“Malaria illness is one sickness that is serious… It causes the body to become hot, sometimes the high temperature leads to convulsion in a child and can lead to the death” **– Male**.*

> *“Some people that lack knowledge or are forgetful. When it is a day to their appointment we call them or even go to their homes to remind them to bring their child to the centre to get immunization and PMC….” Community mobilizer*

Self-efficacy and confidence play a vital role in improving an individual’s capability to take action to improve their health and well-being. Self-efficacy and confidence are crucial components of the capability component of the COM-B model.

> *“I delivered my twins here (referring to the otan-ile health centre)… The twins collected PMC after 10 weeks of the delivery with injection and I have not noticed malaria in them. They write the date we are to return on our card and I bring them on the date……” WDC*

### Opportunity: Social and Environmental Enablers

Opportunities reflected the environmental and social conditions that facilitated or constrained behaviours related to PMC. Participants noted that once a child was eligible, PMC could be accessed at any convenient time, and caregivers felt comfortable seeking clarification from healthcare workers, who also served as the primary source of information on malaria prevention and treatment services. At the community level, WDCs and community health mobilizers disseminated information on the availability of PMC through community forums, churches, and mosques. These actors also engaged hard-to-reach communities and households providing sensitization and education on the benefits of PMC. Women commonly shared information with spouses and neighbours, encouraging PMC uptake. While men described active support, including providing transport fare, accompanying caregivers to facilities, and reminding them of scheduled appointments. Some men also followed up with their spouses after clinic visits, and health workers routinely called caregivers to remind them of upcoming appointments for their children.

> *“Anytime the children need the medicine it is available, we take the children for it and it works well…**Female.***

> *“We educate them …. for them to ensure that they come for SP for their children… for those we don’t see we call them to ask why” **Health worker***

> *“We do go out to inform our neighbors or community members to prevent this sickness (malaria), we ask them to come to oke odo (health center) to collect the medicine (PMC) for their children” **Community mobilizer.***

> *“…. The ways we support our wives….I usually remind my wife to take the child for his appointment and if the child is slightly hot, I will ask her to take the child to the health facility. Sometimes I take them there myself…” **Male**.*

Participants identified long waiting times at health facilities as a key barrier to accessing PMC linked to RI services. Caregivers reported that prolonged delays discouraged attendance, particularly for women who needed to balance childcare with other livelihood responsibilities. These waiting times were attributed to inadequate staffing and the resulting high workload placed on the limited number of healthcare workers.

> ***“****The challenges that face our people… is that people are many, some women may want to go to their market and will be in a haste…But if health workers are many in the health facilities this will make individuals get the health care that they want on time….” **Male**.*

> *“Nursing mothers used to be much more in number than the number of healthcare workers, … So the number of hours that some mothers spend there used to discourage them and they will say they will not come again, ….” **WDC***.

Participants providing suggestions and solutions to the challenges, described how healthcare services could be made more accessible and convenient for the children, and how technology (e.g., mobile apps, text messages) could be used to support adherence to PMC. Some of the suggestions included house to house campaign on the PMC, making PMC available in all government health centres in both communities, more mass media campaign and use of social media such as facebook, WhatsApp to remind mothers of their scheduled appointment.

> *“The way out is firstly government should recruit more staffs…. Secondly, there should be more public enlightenment, the news should be more on air (referring to radio and TV), online on facebook, in the community the more the news is spread, those that have benefitted can also assist us by telling others….” **WDC .***

### Motivation: Perceived Benefits and Threats

Multiple factors were identified as motivating caregivers to access PMC. These included the perceived seriousness of malaria, recognition of its potential to cause severe illness or death, and the perceived benefits of PMC in reducing the malaria burden among young children. Reminders from community mobilizers and trust in healthcare workers further reinforced caregivers’ motivation. Participants consistently described malaria as a common and serious health threat, which encouraged them to prioritise preventive actions to keep their children healthy. Caregivers also noted that a child’s illness disrupted household stability and limited women’s ability to attend to other responsibilities, reinforcing the value they placed on prevention. Spousal support also emerged as an important motivator; some men reported providing financial and emotional support to enable their wives to attend appointments, noting visible improvements in their children’s health as a result. Health workers reported a noticeable decline in malaria cases among children under two years following PMC implementation, reinforcing caregivers’ perceptions of its effectiveness. The WDCs similarly emphasised the observed benefits of PMC and described ongoing efforts to educate mothers and encourage those delivering in traditional birth homes to bring their newborns to health facilities for vaccination and preventive medicines.

> *“I have heard several cases when the child died from a serious illness like malaria. …. By the time they went to the hospital the sickness had escalated and it was too late. Eventually the child died. Serious malaria can kill…” **Health worker.***

> *“Previously malaria was high but now it is reducing, our own facility is in front of a school. Previously they used to bring the children to us that they are vomiting with high temperature, we are not seeing all these again among children, it has reduced…” **Health worker.***

> *“One of the supports I give to my wife… I give her financial support …This also motivates her to come. Since she has been doing all these things there is a lot of changes in my family in terms of malaria, it is no longer common in my family” **Male.***

### Behaviour towards PMC

Following identification and mapping of the targeted behaviour, a workshop with stakeholders was conducted to determine key behaviours to focus on to ensure that children complete their doses for immunization and PMC. Emphases were placed on developing clear, concise, and culturally relevant messages to promote desired behaviors. These messages were included in a strategic behaviour change communication materials (community dialogue flipbook and community dialogue facilitator guide). All messages projected the positioning statement that “PMC is “***the gateway for Child Healthy growth and thriving***”. The materials were subsequently pretested to refine and finetune the messages and also obtain feedback from community members. Figure 2 shows the COM- B model framework for target audience motivation to take action to protect their children:

**Figure 2:**
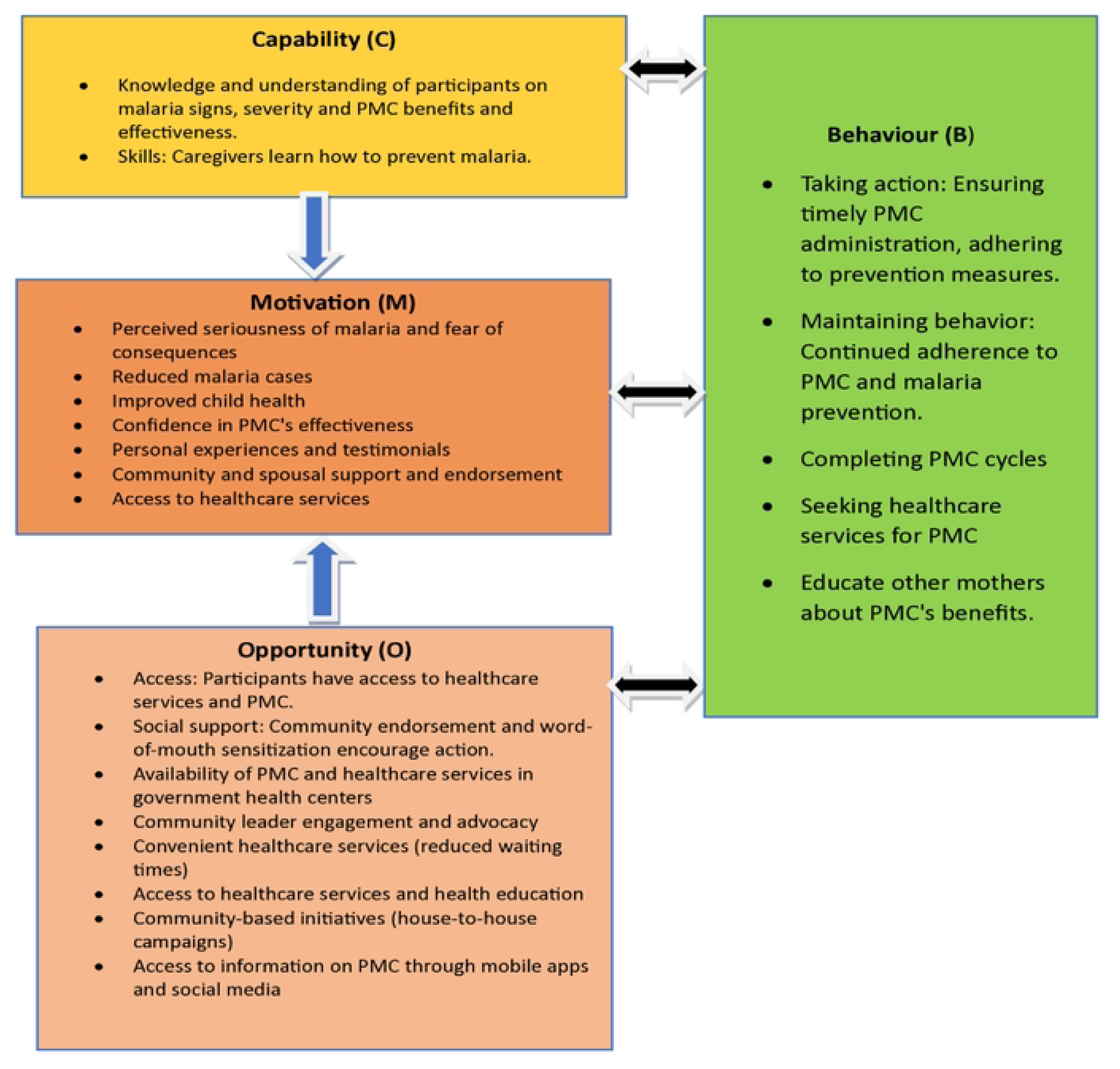
The COM- B model framework for target audience motivation to take action to protect their children from malaria

### SBCC delivery

A total of 81 community mobilisers (20 men and 61 women) were trained and reached 24,920 persons (6,519 men and 18,401 women) across 80 communities in eight LGAs in Osun State between November 2024 and August 2025.

## Discussion

This study demonstrates the practical utility of the COM-B behaviour change model in informing the design of culturally appropriate SBC messages to support uptake of PMC delivered through the RI platform in Osun State, Nigeria. By systematically identifying behavioural drivers and barriers across capability, opportunity, and motivation domains, the study provides evidence on how behavioural science can be operationalised to strengthen malaria prevention interventions in high-burden settings.

Caregivers in this study demonstrated substantial knowledge of malaria symptoms, severity, and consequences, which is required to influence care-seeking behaviour and acceptance of preventive interventions. Recognition of danger signs such as fever, convulsions, anaemia, and lethargy aligned with findings from other sub-Saharan African settings where symptom awareness enhances prompt healthcare utilisation and adherence to malaria interventions[2,3,20]. The observed self-efficacy among caregivers—particularly confidence in adhering to appointment schedules and administering PMC alongside immunisation—reflects the importance of psychological capability in sustaining preventive behaviours. However, forgetting the time to visit the health facility emerged as a persistent capability-related barrier, particularly for monthly PMC dosing. Similar challenges have been documented for RI attendance and other malaria chemoprevention strategies[21,22]. This underscores the need for SBC strategies that move beyond information provision to include practical memory aids such as reminder cards, mobile phone prompts, and follow-up by community mobilisers.

Opportunity-related factors played a critical role in shaping PMC uptake. Social support—especially spousal involvement—emerged as an enabler. Men’s financial, logistical, and emotional support not only facilitated clinic attendance but also reinforced women’s motivation to sustain preventive practices. This finding aligns with growing evidence that male engagement improves uptake of maternal and child health interventions, including malaria prevention[23]. Community mobilisers, ward development committees, and trusted health workers were key social influencers who enhanced access to information and services. Their role in community sensitisation, appointment reminders, and trust-building reflects the value of community-embedded delivery approaches for new or pilot interventions such as PMC. Conversely, long waiting times at health facilities were identified as a major opportunity-related barrier, driven largely by human resource constraints. Similar findings have been reported across Nigeria and other low-resource settings, where inadequate staffing undermines immunisation performance and discourages repeat attendance[24,25]. Given that PMC is integrated into the RI platform, addressing systemic bottlenecks affecting immunisation services is essential for optimising PMC uptake. Strengthening health workforce capacity and improving service organisation would therefore have dual benefits for RI and PMC.

Motivation to access PMC could be influenced by caregivers’ perception of malaria as a serious and potentially fatal disease, particularly among young children. Fear of severe outcomes and previous experiences with malaria-related illness or death reinforced preventive action, consistent with findings from studies applying the Health Belief Model to malaria prevention behaviours[26–28]. Importantly, caregivers and health workers reported observable reductions in malaria episodes following PMC implementation, which strengthened confidence in the intervention and reinforced continued participation. Such experiential reinforcement is a powerful motivator and supports sustained behaviour change beyond initial adoption. Fathers’ testimonies of reduced household malaria burden further highlight how visible health gains can transform motivation into advocacy within families and communities. Nevertheless, a lack of perceived urgency among some caregivers remained a demotivating factor, particularly when children appeared healthy. This finding mirrors challenges documented for RI drop-out in the second year of life and underscores the importance of SBC messages that emphasise prevention even in the absence of illness[29].

Applying the COM-B model enabled a structured, theory-driven approach to message development by clearly linking identified barriers and enablers to targeted SBC strategies. Capability gaps informed simplified, locally relevant messages on malaria risk and PMC benefits; opportunity constraints guided the promotion of social support and community-based reminders; and motivational drivers shaped testimonials and emotional appeals centred on child survival and wellbeing. This study adds to the growing body of evidence supporting the application of COM-B across diverse health contexts, including infectious disease prevention and service uptake[18,19,30,31]. By translating qualitative insights into actionable SBC materials—such as community dialogue flipbooks and facilitator guides—the study demonstrates how behavioural theory can be embedded into routine programme design.

## Strengths and limitations

A key strength of this work is the inclusion of multiple stakeholder groups, enabling a holistic understanding of behavioural influences across household, community, and health-system levels.

The participatory approach to SBC message development further enhanced contextual relevance and acceptability.

However, findings are based on qualitative data from two LGAs and may not be fully generalisable to all settings in Nigeria. Other limitations include social desirability bias, recall bias and confirmation bias as the researchers unconsciously interpret data to support expected COM-B outcomes. Translation bias with potential loss of meaning or nuance during translation is also a limitation in the study.

Additionally, while the study describes SBC reach, it does not quantitatively assess changes in PMC uptake attributable to the intervention. Future research should incorporate longitudinal and mixed-methods evaluations to measure behavioural and epidemiological outcomes.

## Conclusion

The COM-B model provided an effective framework for identifying the capability, opportunities, motivation and factors influencing uptake of PMC. Key enablers included knowledge, health systems accessibility, spousal support and perceived benefits. Addressing barriers such as forgetting the time to visit the health facility, long wait times and low perceived urgency —while reinforcing enablers including spousal support, trusted health workers, and visible health gains—will be critical for sustaining PMC uptake and maximising its contribution to malaria control in Nigeria and similar high-burden settings.

## Data Availability

The data generated and/or analysed during the current study are not publicly available due to analysis being underway for subsequent publications. Once study is completed data will be made publicly available upon request

## Acknowledgements

We wish to express our profound gratitude to the caregivers, fathers, local leaders, health workers and community mobilizers who participated in this work. We also appreciate the researchers and State Ministry of Health staff who provided technical support in the conduct of this work.

## Contributions

All authors made a significant contribution to the work and took part in drafting, revising or critically reviewing the article and gave final approval of the version to be published.

